# MAUDGAN: Motion Artifact Unsupervised Disentanglement Generative Adversarial Network of Multicenter MRI Data with Different Brain tumors

**DOI:** 10.1101/2023.03.06.23285299

**Authors:** Mojtaba Safari, Ali Fatemi, Louis Archambault

## Abstract

**Purpose:** This study proposed a novel retrospective motion reduction method named motion artifact unsupervised disentanglement generative adversarial network (MAUDGAN) that reduces the motion artifacts from brain images with tumors and metastases. The MAUDGAN was trained using a mutlimodal multicenter 3D T1-Gd and T2-fluid attenuated inversion recovery MRI images.

**Approach:** The motion artifact with different artifact levels were simulated in *k*-space for the 3D T1-Gd MRI images. The MAUDGAN consisted of two generators, two discriminators and two feature extractor networks constructed using the residual blocks. The generators map the images from content space to artifact space and vice-versa. On the other hand, the discriminators attempted to discriminate the content codes to learn the motion-free and motion-corrupted content spaces.

**Results:** We compared the MAUDGAN with the CycleGAN and Pix2pix-GAN. Qualitatively, the MAUDGAN could remove the motion with the highest level of soft-tissue contrasts without adding spatial and frequency distortions. Quantitatively, we reported six metrics including normalized mean squared error (NMSE), structural similarity index (SSIM), multi-scale structural similarity index (MS-SSIM), peak signal-to-noise ratio (PSNR), visual information fidelity (VIF), and multi-scale gradient magnitude similarity deviation (MS-GMSD). The MAUDGAN got the lowest NMSE and MS-GMSD. On average, the proposed MAUDGAN reconstructed motion-free images with the highest SSIM, PSNR, and VIF values and comparable MS-SSIM values.

**Conclusions:** The MAUDGAN can disentangle motion artifacts from the 3D T1-Gd dataset under a multimodal framework. The motion reduction will improve automatic and manual post-processing algorithms including auto-segmentations, registrations, and contouring for guided therapies such as radiotherapy and surgery.

## 1 Introduction

Magnetic resonance imaging (MRI) with different sequences provides excellent soft tissue contrast for diagnosis and treatment planning. However, high MRI acquisition time limits the quality of high-resolution images^1^ because of the increased probability of patient motion. Involuntary and voluntary subject motions during data acquisition cause image blurring and ghosting along the phase-encoding direction. The prevalence of motion artifacts is high for infants and patients with acute distress.^2^

To tackle motion artifacts, retrospective motion correction (RMC) and prospective motion correction (PMC) methods were developed. PMC approaches modify the gradient magnetic fields using the imaged object positions that are tracked during imaging to maintain a constant relationship between imaged object and imaged volume.^3, 4^ PMC can maintain a uniform *k*-space sampling density, which avoids Nyquist violation, and compensate for spin-history effects.^5^ However, PMC methods require additional hardware and complicated pulse sequences that increase the imaging time. On the other hand, RMC methods are post-processing approaches, and do not require additional hardware and pulse sequence modifications during imaging. Traditional RMC methods, such as auto-focusing, attempt to optimize image quality metrics like entropy and gradient,^6^ iterative methods to estimate motion trajectory,^7^ compressed-sensing theory,^8^ and modified imaging sequences.^9^ They are either limited to 2D imaging methods or require raw *k*-space data that are not widely available. In addition, these methods are computationally expensive.

Recently, deep learning techniques, in particular, convolutional neural networks (CNNs) have been used to quantify^10, 11^ and reduce^12, 13^ MRI motion artifact retrospectively. These models learn the task through a supervised framework using the simulating motion artifacts. Unpaired deep learning models attempted to use data without the motion artifact as a ground truth to reduce the artifacts from MRI with the same imaging sequence.^14^

This study aimed to address the problem in a more practical setting where one motion-free MRI modality removes artifacts from the motion-corrupted images acquired with different MRI imaging sequences. This study reformulated MRI motion artifacts as an unsupervised disentanglement problem. Thus, we introduced a novel motion artifact unsupervised disentanglement generative adversarial network (MAUDGAN). The novel MAUDGAN was applied to reduce the motion of 3D T1-Gd MRI sequences using motion-free T2-fluid attenuated inversion recovery (FLAIR) sequences for the patients with different brain cancers metastasis. This study used a multicenter dataset to improve the MUADGAN’s generalization.

This study leverages an inductive bias^15^ that the MAUDGAN learn to disentangle motion artifacts from motion-free contents by comparing 3D T1-Gd MRI sequences (typically with motion artifacts) with motion-free T2-FLAIR (Figure 1) in the latent space.

**Fig 1:**
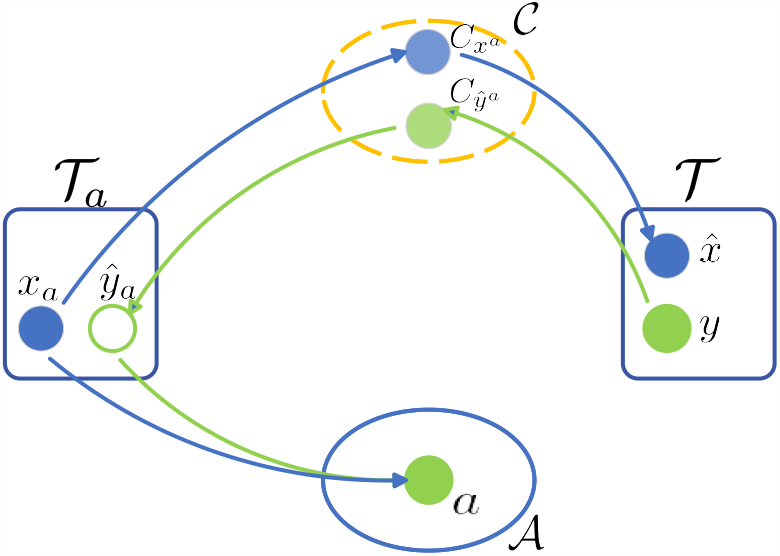
Content and artifact components of 3D T1-Gd MRI (*x*_*a*_) in the motion-corrupted space 𝒯_*a*_ and T2-FLAIR in artifact-free space 𝒯 are mapped to the content space *𝒞* and artifact space *𝒜*, respectively. MAUDGAN maps the data in 𝒯_*a*_ space to 𝒯 space 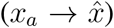 shown by blue arrows. Conversely, MADuGAN learns to map from 𝒯 space to 𝒯_*a*_ space (*y* → *ŷ*_*a*_) shown by green arrows.

The MAUDGAN consists of U-net^16^ generators to perform different forms of image translations including motion artifact reduction and synthesis. Discriminators were used to distinguish between the motion-free and the motion-corrupted MRI sequences in the latent spaces. To our knowledge, MAUDGAN is the first study in multi-modal anatomical MRI motion artifact reduction.

The rest of this paper is as follows: Section 2 explains the dataset and motion simulation steps. Section 3 gives a detail about the MAUDGAN architecture and loss functions. Results and comparisons with two generative models are illustrated in Section 4. Finally, Sections 6 and 5 discuss the significance of the MAUDGAN and its possible use in the context of diagnosis and therapy.

## 2 Material

### 2.1 Dataset

We used a publicly available multicenter medical GLIS-RT dataset from the Cancer Imaging Archive^17^ consisting of 230 patients (100 males and 130 females). All patients with different brain tumor types underwent 3D T1-Gd, 2D T2-FLAIR MRI sequences, and a CT scan under different imaging protocols. The brain tumor types were glioblastoma (GBM - 198 cases), anaplastic astrocytoma (AAC - 23 cases), astrocytoma (AC - 5 cases), anaplastic oligodendroglioma (AODG - 2 cases), and oligodendroglioma (ODG - 2 case). We used 80% (11246 image slices) and 20% (2276 image slices) of data for training and testing our method, respectively.

The median of the T2-FLAIR and 3D T1-Gd images’ resolution was 1.1×1.1×5 mm^3^ (standard deviation 0.53 × 0.53 × 0.87 mm^3^) and 0.94 × 0.94 × 1. mm^3^ (standard deviation 0.24 × 0.24 × 1.21 mm^3^), respectively. The T2-FLAIR imaging parameters were (median ± std); TE = 119 ± 64.06 ms, TR = 9000 ± 936.20 ms, TI = 2500 ± 174.02 ms, and flip angle = 150° ± 13.56°. Those parameters for T1-Gd were (median ± std); TE = 2.98 ± 3.86 ms, TR = 2200 ± 1031.76 ms, TI = 900 ± 235.50 ms, and flip angle = 9° ± 5.45° About 30% of data were acquired using MRI scanners with B_0_ of 1.5 T and the others were acquired using 3T scanners. Out of 230 cases, 55 cases were obtained using GE MRI scanners and the rest were obtained using Siemens MRI scanners.

Finally, we evaluated the MAUDGAN performance on anonymized clinical data with real motion artifacts. This retrospective single-centre study was approved by the institutional review board, and the requirement for written informed consent was waived.

### 2.2 Motion simulation

The head motion was simulated in the Fourier domain (*k*-space), and the motion-corrupted data was generated after the inverse discrete Fourier transform. We adapted the piecewise constant motion simulation approach with a low computation burden because it provides a similar generalization than to the complex motion simulation techniques.^13^ Moreover, the generated motion artifacts were similar to the real motion artifacts.^13^

We assumed the phase encoding interval was much faster than the head motion. Thus, the same motion parameters could be used at each phase encoding direction (Figure 2). The *k*-space lines within the randomly selected slabs were translated in the phase encoding direction. However, the middle of the *k*-space that corresponds to the low-frequency content of the MRI images was excluded in the motion artifact simulation process, shown as a forbidden region in Figure 2. Our motion simulation method could successfully model the ghosting of the bright fat tissue, due to the motion artifact, to the background around the skull, which is common in structural MRI images.^18^

**Fig 2:**
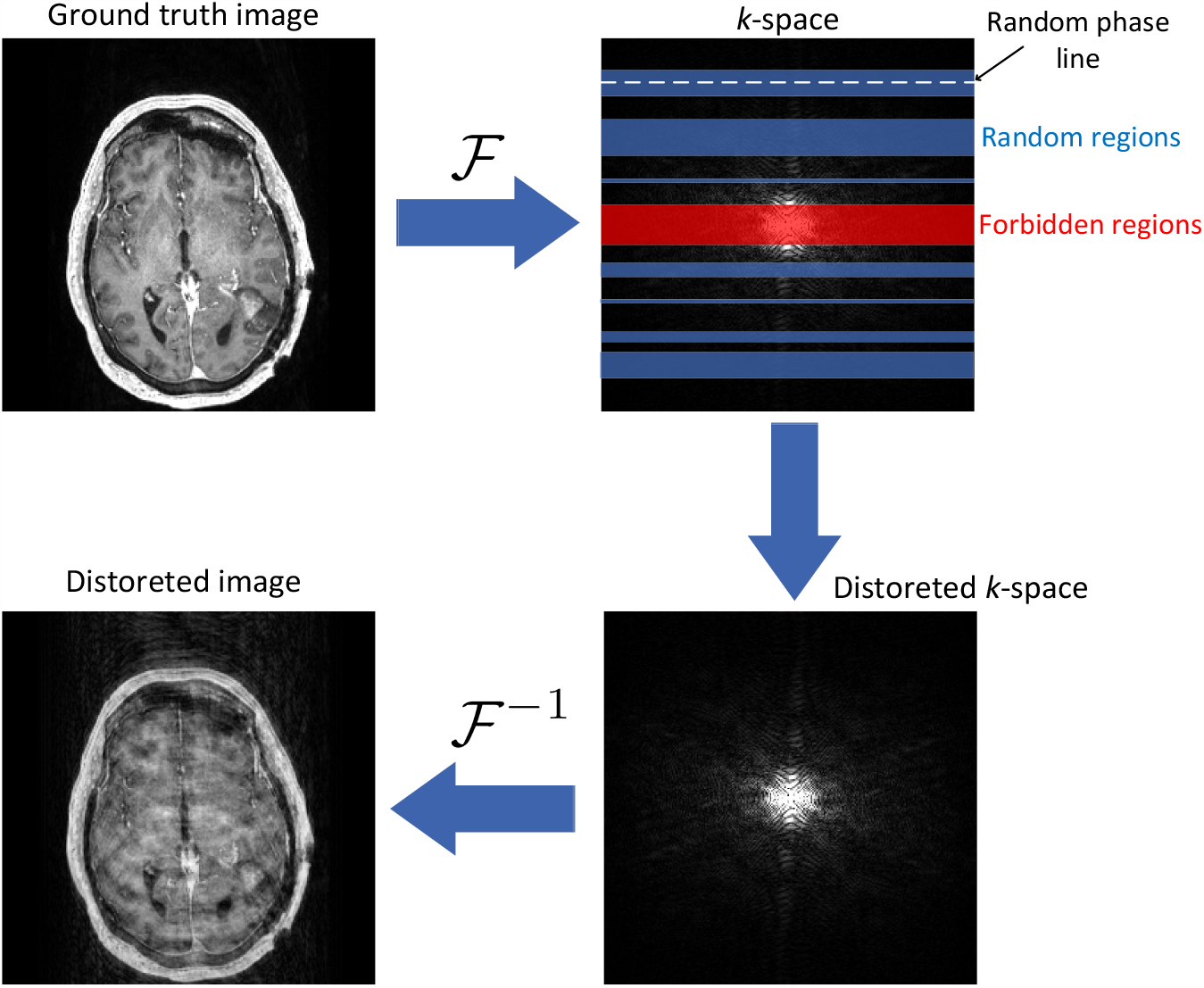
The motion simulation process. After choosing the phase encoding direction, several random *k*-space regions were selected. The randomly selected *k*-space lines were randomly translated within the random regions.

## 3 Method

We denote *𝒯*_*a*_ and *𝒯* as the motion-corrupted image and the motion-free image spaces, respectively. The paired and unpaired motion reduction process is formalized as a *ℳ* = *{*(*x*_*a*_, *x*) | *x*_*c*_ ∈ *𝒯* _*a*_, *x* ∈ *𝒯, f* (*x*_*a*_) = *x}* where *x*_*a*_ and *x* were the motion-corrupted and motion-free single MRI image sequence and *f* : *𝒯*_*a*_ → *𝒯*.^14, 19^ However, we assumed there is no paired or unpaired dataset of a single modality available to disentangle motion artifacts. Instead, another MRI image sequence, T2-FLAIR, was employed to disentangle the motion artifact of the T1-Gd MRI sequence, which is more practical in clinical settings. Thus, the MAUDGAN is formalized as *ℳ* = *{*(*x*_*a*_, *y*) | *x*_*a*_ ∈ *𝒯* _*a*_, *y* ∈ *𝒯, f* (*x*_*a*_) = *x, g*(*x*_*a*_, *y*) = *y*_*a*_*}* where *f* : *𝒯*_*a*_ → *𝒯* and *g* : *𝒯* → *𝒯* _*a*_ are the encoding into a content space *𝒞* and artifact space *𝒜*. Also, *x*_*a*_ and *y* are motion-corrupted T1-Gd and the motion-free T2-FLAIR MRI images. After training the MAUDGAN, the image data in the content space will be free of motion artifacts. In contrast, the motion-corrupted T2-FLAIR could be generated using the learned motion artifact model.

### 3.1 MAUDGAN

The MAUDGAN consists of two generators 𝒯 : 𝒯 _*a*_ → *𝒯* and *𝒢* : *𝒯* → *𝒯* _*a*_ to map from motion-corrupted space to motion-free space and vice-versa (Figure 3). In addition, two networks 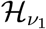 and 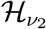 were also employed to extract features of the images before feeding them to the generators.

**Fig 3:**
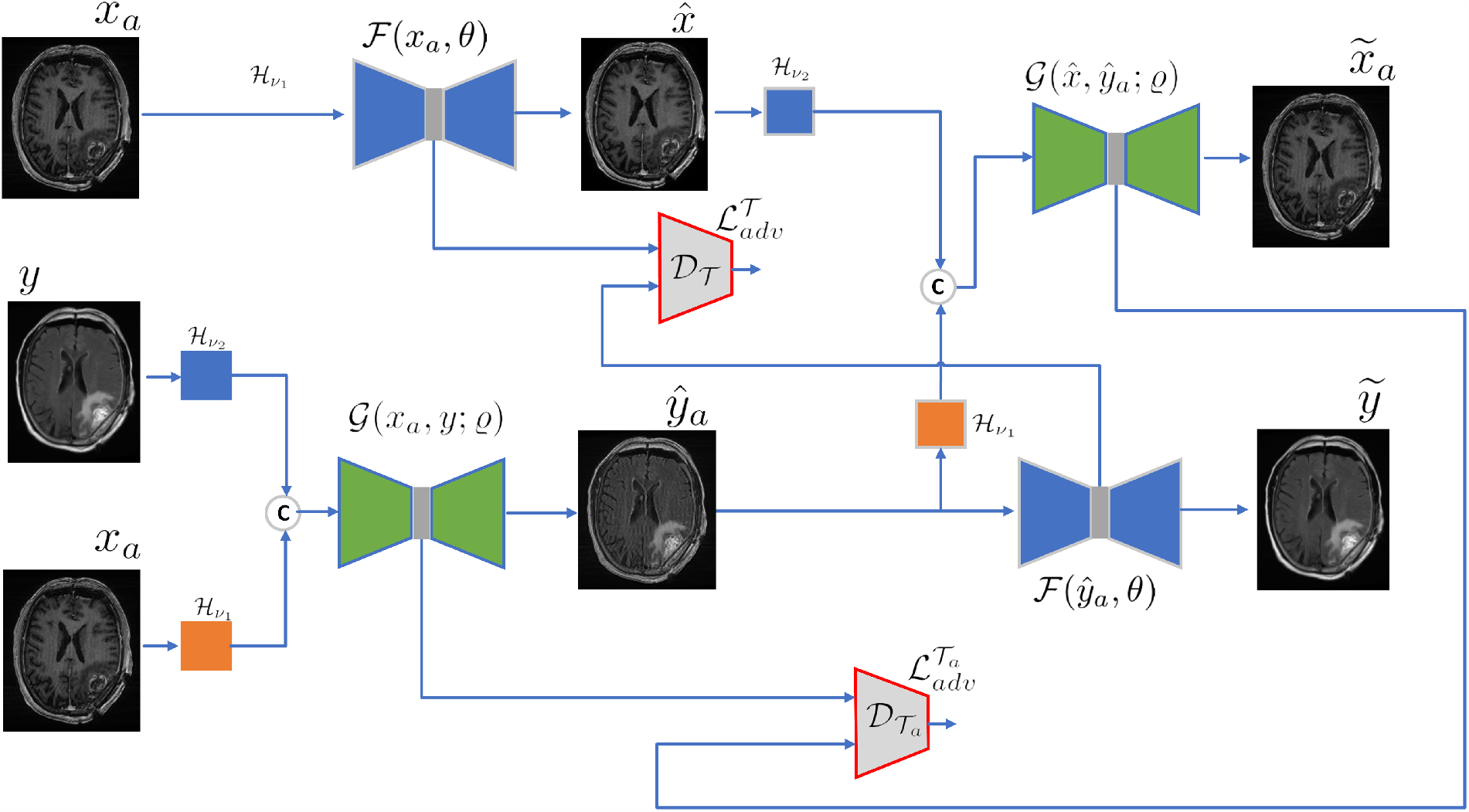
The proposed MAUDGAN is illustrated. The Generators *ℱ* learns disentanglement while the *𝒢* learns to generate motion-corrupted images from motion-free images.

Given multimodal MRI images T1-Gd *x*_*a*_ ∈ *𝒯* _*a*_ and T2-FLAIR *y* ∈ *𝒯*, the training steps were as follows:

1. *ℱ* maps the motion-corrupted T1-Gd *x*_*a*_ to motion-free space 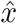,

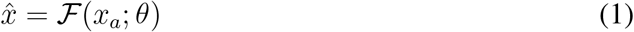
2. *𝒢* maps the motion-free space T2-FLAIR *y* to the motion-corrupted space *ŷ*_*a*_,

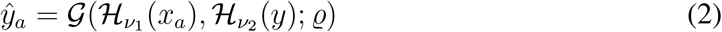
3. trained *ℱ* in step 1 was used to recover motion-free T2-FLAIR 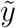 from motion-corrupted *ŷ*_*a*_ simulated in step 2,

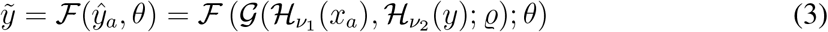
4. trained *𝒢* in step 2 was used to recover motion corrupted T1-Gd 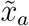 from motion-free 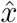 and motion-corrupted *ŷ*_*a*_ simulated in step 1 and 2,

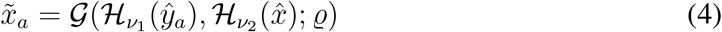

### 3.2 Learning

The MAUDGAN attempts to train generators in an adversarial scenario to achieve motion artifact disentanglement. Thus, the MAUDGAN employed loss functions to remove motion artifacts from T1-Gd using content information of T2-FLAIR as given in (1)-(4). The MAUDGAN employs four loss functions including two adversarial losses 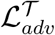 and 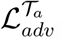, reconstruction loss *ℒ*_*rec*_, and artefact consistency loss *ℒ*_*arti*_. The cost function is formalized as the weighted sum of the losses,

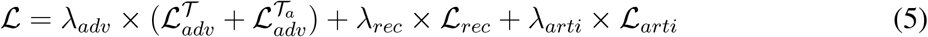

where *λ*_*adv*_, *λ*_*rec*_, and *λ*_*arti*_ are the hyper-parameters controlling the importance of each term.

#### 3.2.1 Adversarial loss

The MAUDGAN was trained to map from motion-corrupted space to motion-free space as given in (1) and (3) and vice versa as given in (2) and (4). Learning those two tasks are important to disentangle motion artifact from the image content. As the MAUDGAN is trained on multimodal MRI sequences, regression losses like *ℒ*_1_ and *ℒ*_2_ could not be employed due to the domain difference between T2-FLAIR and T1-Gd MRI images. Therefore, the adversarial learning technique,^20^ introduced *𝒟*_*T*_ and 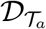 discriminators, was employed to regularize the plausibility between motion-corrected and motion-free images using 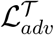 loss and between motion-corrupted and motion-simulated images using 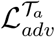 loss. Thus, the MAUDGAN is trained to fool the discriminators, so they could not determine whether the motion was generated or real. The adversarial losses are as follows;

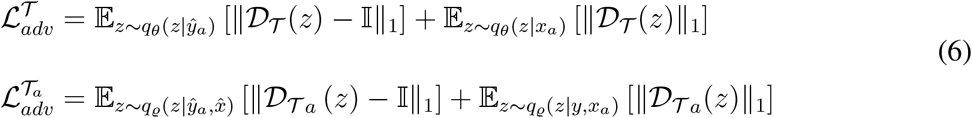

where *z* is the latent variable generators, *𝒟*_*T*_ and 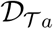 are the discriminators to distinguish between motion-corrupted and motion-free content data sampled from *𝒯* and *𝒯*_*a*_ domains, respectively. 𝕀 is an unit matrix with a size *M* × *M*, where *M* is substantially smaller than the image dimension size, that matched the discriminators’ output.

#### 3.2.2 Reconstruction loss

Despite motion artifact disentanglement, the whole process needed to be lossless. In other words, the MAUDGAN was required to recover the original motion-corrupted T1-Gd 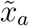 from motion-corrected 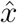 and to recover motion-free T2-FLAIR 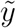 from motion-simulated *ŷ*_*a*_. Therefore, two reconstruction losses given in (7) were used to encourage the MAUDGAN to preserve the information.

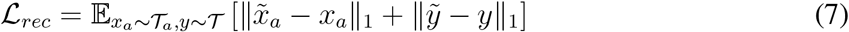

where 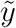 and 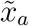 shown in Figure 3 are given in (3) and (4). We adapted the *ℒ*_1_ loss rather that than the *ℒ*_2_ to generate sharper images.^21^

#### 3.2.3 Artifact consistency loss

Adversarial losses encouraged the content of generated motion-corrupted *ŷ*_*a*_ and motion-free 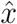 images to be indistinguishable from T1-Gd *x*_*a*_ and T2-FLAIR *y* images, respectively. However, the discriminators lose the spatial resolution. To preserve the spatial resolution, *ℒ*_1_ and *ℒ*_2_ could be used. But, due to the domain difference between T1-Gd and T2-FLAIR, direct use of losses would transfer the images’ domain. We proposed artifact loss *ℒ*_*artif*_ given in (8) to induce motion artifacts to the motion-corrected 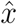 images. Thus, *ℒ*_*artif*_ conflicts with adversarial losses and comprises the overall learning process.

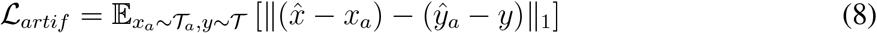

Equation (8) encourages the difference between *x*_*a*_ and 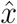 to be similar to *y* and *ŷ*_*a*_. Unlike a direct minimization by *ℒ*_1_ that would cause an image domain translation, *ℒ*_*artif*_ requires the 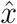 and *x*_*a*_ to be anatomically close rather be exactly close to preserve structural information.

### 3.3 Network architecture

The MAUDGAN network generator is illustrated in Figure 4-(a). The generator employed residual blocks^22^ (Figure 4-(b)) for a better generalization than convolution blocks without skip connection. To improve the generators’ performance,^23^ the convolution layers were used to down-sample the data in the encoder part of the generator. However, the decoder part of the generator employed the up-sampling layers rather than the transpose convolution layers to preserve the image edge information and avoid the checkerboard effect.^24^

**Fig 4:**
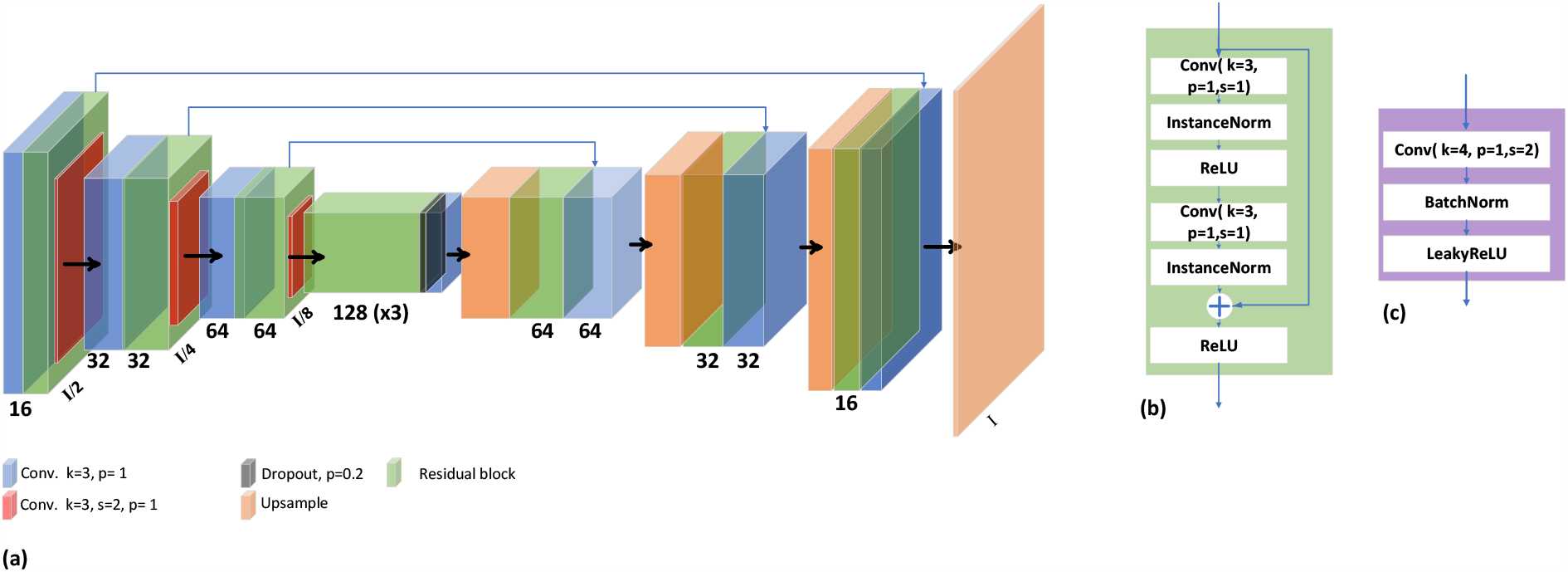
The Generator with the blocks used to construct discriminator and 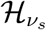 are illustrated.

The discriminator consists of four residual blocks (Figure 4-(b)) and down-sampling blocks. Finally, the discriminators were constructed by four convolution blocks shown in Figure 4-(c) and the final layer with one convolution layer. The feature extractors (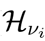 for *i* ∈ *{*1, 2*}*) were constructed using five residual blocks (Figure 4-(b)).

We implemented the MAUDGAN under the `PyTorch 1.12.0`^1^ deep learning framework using two NVIDIA GPUs RTX 3090. The batch size, optimizer, and the learning rate were 6, RAdam,^25^ and 2 × 10^−4^. We trained the network using hyper-parameters *λ*_*rec*_ = 10, *λ*_*adv*_ = 5, and *λ*_*artif*_ = 50.

## 4 Results

To our knowledge the MAUDGAN is the first network that employs the multi-modal MRI images to reduce MRI motion artifacts. Thus, we could only compare the MAUDGAN with two wellknown unsupervised image-to-image translation approaches including CycleGAN^26^ and Pix2pix.^21^ The original implementations of the CycleGAN and Pix2pix were used^2^ to compare the results.

The supervised methods like U-Net^27^ were excluded since the ground truth targets were unavailable, and the domain shifts between the multi-modal images transfer the domain of the input motion-corrupted 3D T1-Gd images to the motion-free T2-FLAIR dataset. We compared the MAUDGAN with those networks for different motion artifact levels. Finally, we evaluated the performance of the MAUDAN to remove real motion artifacts from the patients with head & neck cancer.

Motion simulated dataset allowed us to perform qualitative and quantitative comparisons. We report six quantitative metrics including normalized mean squared error (NMSE), structural similarity index (SSIM),^28^ multi-scale structural similarity index (MS-SSIM),^29^ peak signal-to-noise ratio, visual information fidelity (VIF),^30^ and multi-scale gradient magnitude similarity deviation (MS-GMSD).^31^ The higher metric values are better regarding motion artifact reduction and distortion levels except with the NMSE and MS-GMSD metrics.

Qualitative comparisons are illustrated in Figure 5 for different motion levels. Qualitatively, the Pix2Pix method had the lowest performance in preserving the MRI soft tissue contrast. CycleGAN reduced soft-tissue contrasts, smeared out the signal intensity, and unrealistically elevated the skull signals. MAUDGAN remove motion artifact with better soft tissue contrast and realistic skull signal intensity.

**Fig 5:**
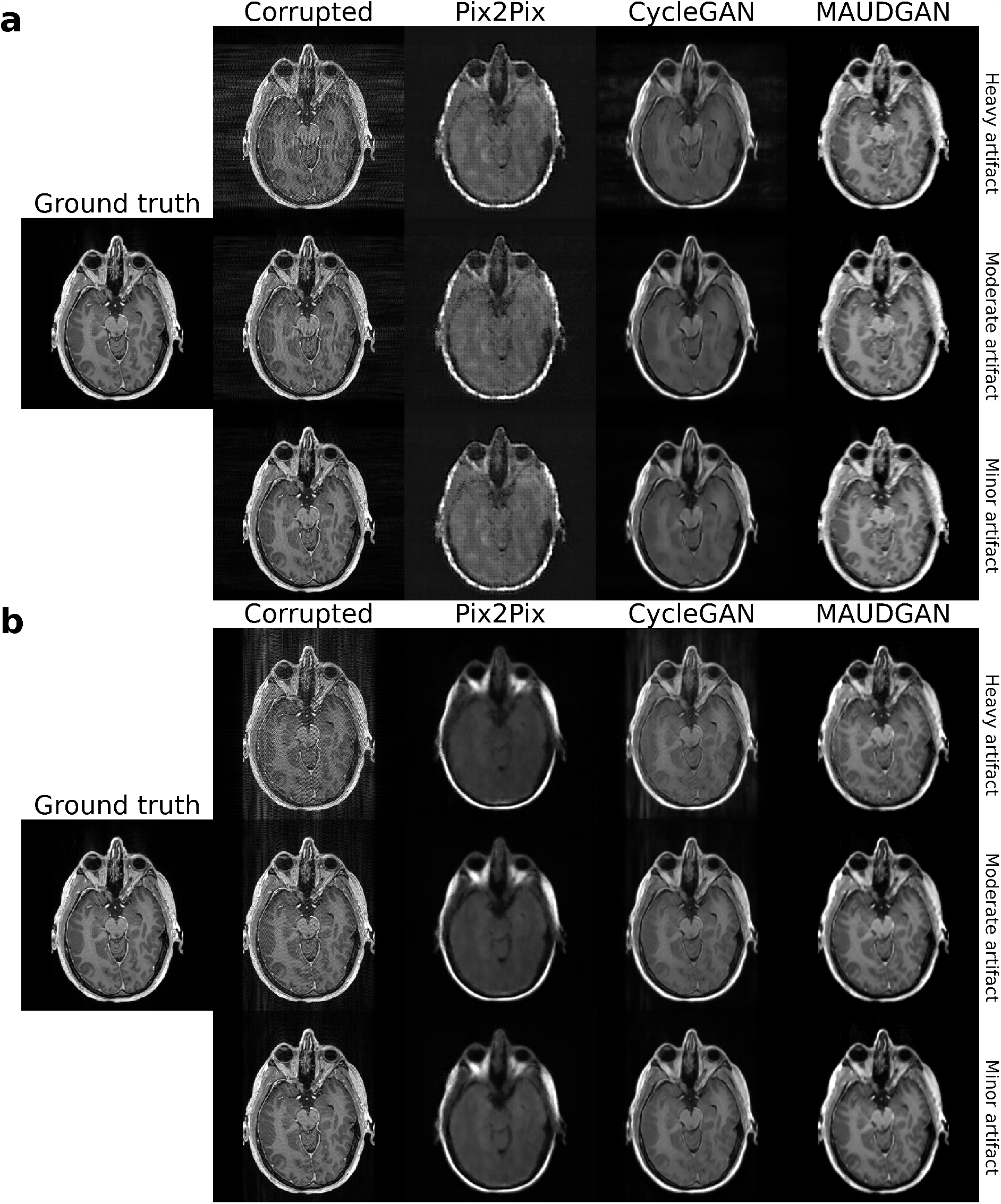
Visual comparisons of the motion-reduction methods on the motion-simulated data. The simulated motion artifact was added along the row in (a) and column in (b). The heavy, moderate, and minor motion simulation data and the motion-corrected results are from top to bottom rows.

In addition, CycleGAN generated images with high signal intensity voxels mimicking the false tumors (see Figure 6). The false tumors were generated might be attributed to the wrong sampling from data manifolds. Those false tumors differ from water droplet-like artifacts^32^ cause by the normalization layers. Especially, the false tumor shown in Figure 6b is similar to the post-surgery cases.

**Fig 6:**
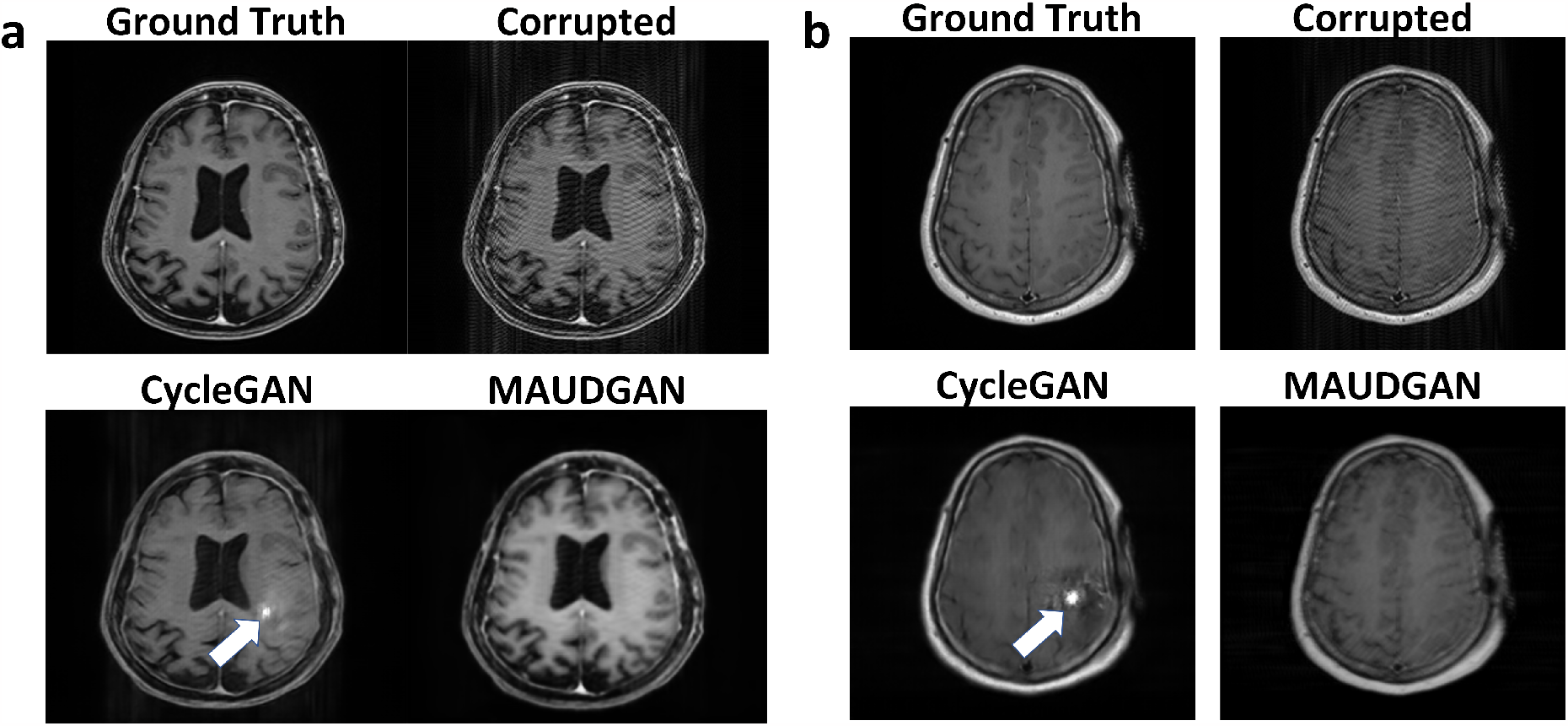
The white arrows illustrate the false tumors generated by the CycleGAN dataset.

The quantitative metrics evaluating the motion-corrected image contrast, image distortion level, and structure and texture similarity to the ground truth data are illustrated in Figure 7. The MAUDGAN with the lowest NMSE and the highest PSNR values indicates the removing the motion artifact with small spatial distortion. However, NMSE and PSNR tend to favor smoothness. The MS-SSIM and SSIM were reported to evaluate the structural similarity of the motion-corrected images and the ground truth. Higher MS-SSIM and SSIM indicate better similarity. Our method got better SSIM values and comparable MS-SSIM values for different distortion levels. The MAUDGAN with the highest value of VIF could preserve more information than the other utilized methods. Finally, to evaluate the image gradient, which is related to image contrast, the MS-GMSD was reported for different distortion levels. Lower MS-GMSD indicates a smaller deviation between the gradients of motion-corrected and ground truth data. The MAUDGAN with smaller MS-GMSD could preserve more, say soft-tissue, the contrast of the ground truth data.

**Fig 7:**
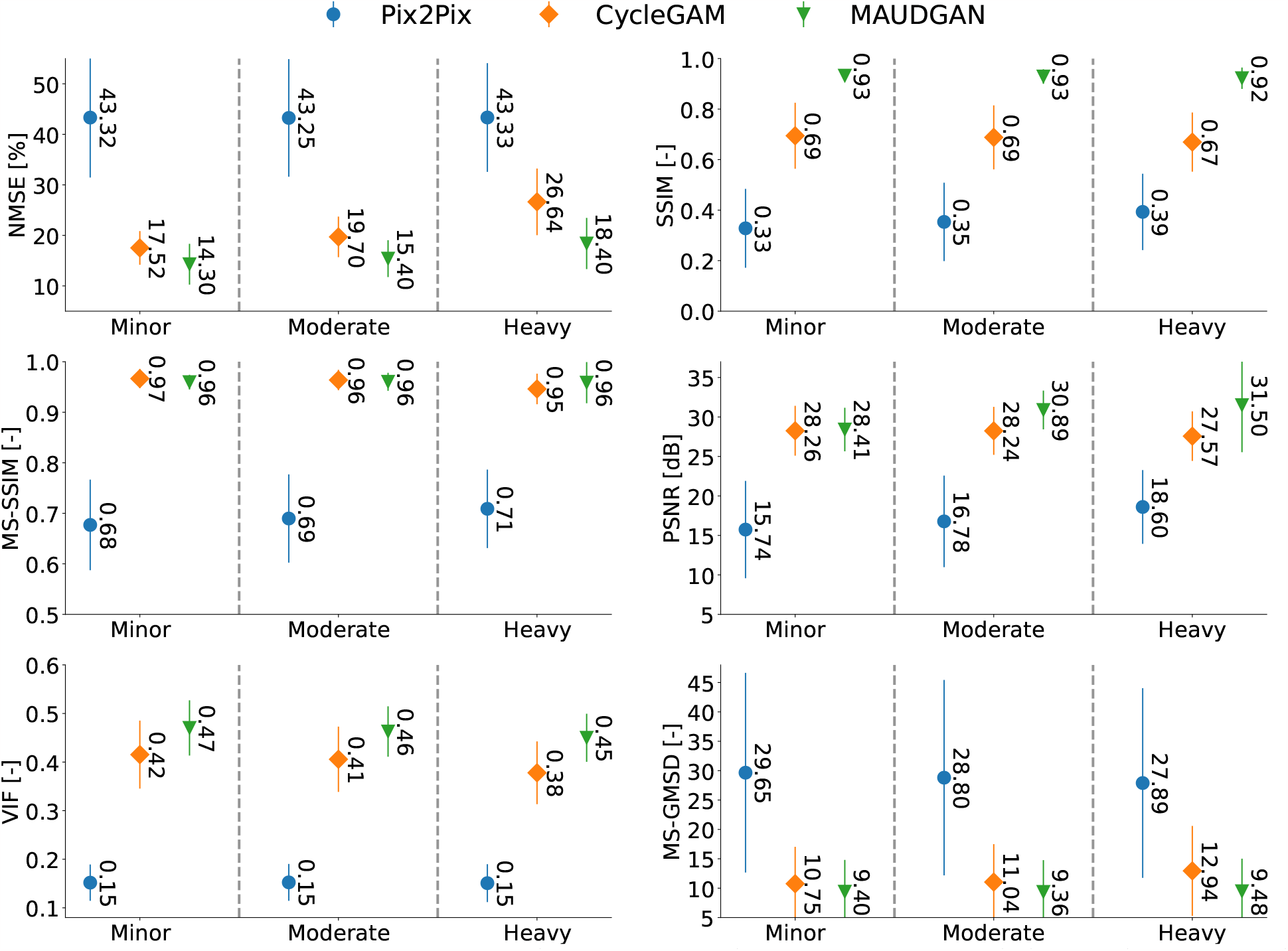
Quantitative metrics to evaluate the quality of the motion-corrected data. The proposed MAUDGAN, Pix2Pix, and CycleGAN were evaluated on three motion distortion levels heavy, moderate, and minor.

We tested the MAUDGAN model on the data with real data with motion artifacts. The data were extracted anonymized from the PACS system. The real artifact was reduced using the MAUDGAN as shown in Figure 8.

**Fig 8:**
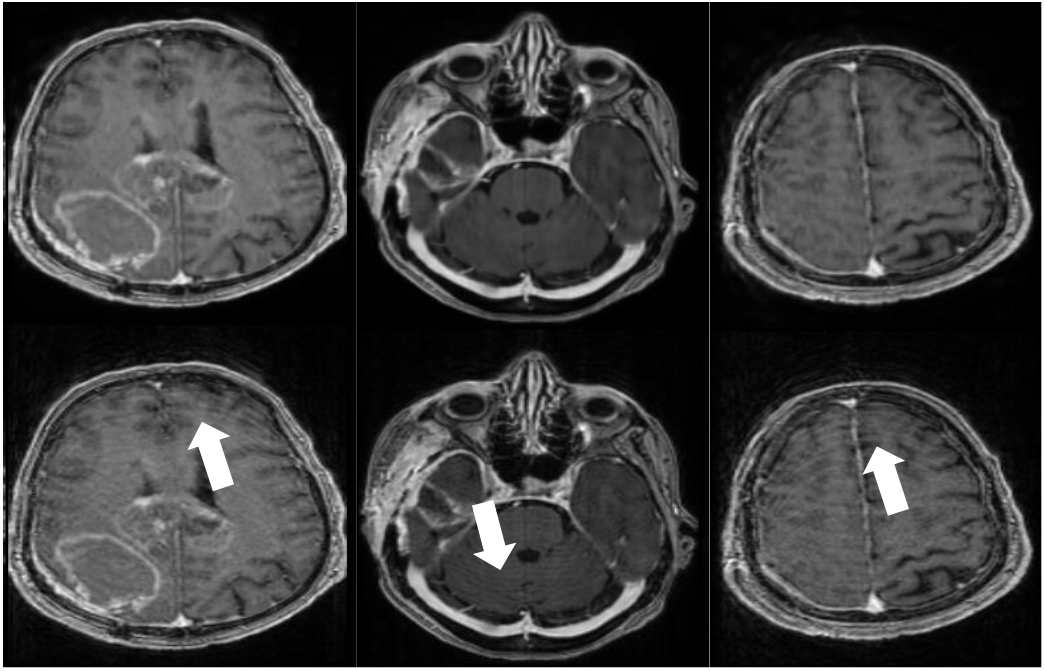
The anonymized data with real motion artifacts were exported from the PACS system to evaluate the MAUDGAN model to remove the real motion artifacts. The first row is the data with real artifact, and the second row illustrates the data after motion reduction. The arrows indicate the motion artifact.

## 5 Discussion

This study aimed to reduce 3D T1-Gd motion artifacts using T2-FLAIR images. 3D T1-Gd images with high acquisition times are more likely to corrupt with the motion artifact.^2^ In addition, the high-resolution images’ quality acquired with the high B0 magnetic fields is limited due to the motion artifact, which the PMC methods could partially remove the motion artifacts.^1^ Motion artifacts reduce the image quality reducing the performance of manual and automatic post-processing approaches like tumor and organ at risks auto-segmentation.^33, 34^ This study introduced MAUDGAN to tackle motion reduction as a disentanglement problem. The multi-center dataset with different brain tumors and metastases was used to train the MAUDGAN, which is expected to improve its generalization. Our qualitative and quantitative comparisons with two well-known GAN methods indicate that the MAUDGAN could disentangle the motion artifact using T2-FLAIR with a lower spatial distortion and a better spatial contrast.

The MAUDGAN was qualitatively compared with generative models CycleGAN and Pix2pix. The MAUDGAN could preserve better soft-tissue contrast (see Figure 5). The Pix2pix approach did not preserve soft-tissue contrast, which might because this method was proposed to work under the paired framework which is different from the theory of this study. On the other hand, the CycleGAN smeared out the MRI soft-tissue contrast, which was better than the pix2pix. Finally, the MAUDGAN reduced the motion artifact with better soft-tissue contrast.

When a network is trained on datasets with tumors, it is crucial that the network to be robust against spatial distortions because those distortions could be misinterpreted as a tumor. The MAUDGAN was free of spatial distortion, while the CycleGAN added spatial distortions (see Figure 6). The added spatial distortions were similar to the brain tumor of the patient with edema and after tumor resection as illustrated in Figure 6(a) and (b), respectively.

The quantitative comparisons shown in Figure 7 between the motion-free ground truth dataset and motion-corrected reconstructed by the CycleGAN, Pix2pix, and MAUDGAN suggest that the MAUDGAN-generated images were more distortion-free with a lower NMSE and a higher PSNR. In addition, MAUDGAN with the higher SSIM, MS-SSIM, and VIF and lower gradient deviations (MS-GMSD) generated more similar to the ground truth dataset.

To the best of our knowledge, this is the first study reporting on the feasibility of an approach enabling to disentangle motion of 3D T1-Gd using T2-FLAIR. The dataset contains different brain tumors and metastases, which are enhanced differently on the different MRI sequences. Thus, we did not use motion-free images of other patients, which need to be exported from PACS. This way, the dataset of the patients without motion artifacts remain in the clinical system. Moreover, we can use all the data to train the network, which is more than training under an unpaired scenario since we do not need to export the same number of patients’ data without motion artifacts.

This study is more challenging compared with the unpaired studies^14, 35^ because the data space domain of 3D T1-Gd differs from T2-FLAIR. Thus, the MAUDGAN must be robust to the domain shift between datasets. Due to the MAUDGAN’s robustness, it could employ other image modalities like the T1-w dataset instead of T2-FLAIR. Thus, the MAUDGAN applies to other available MRI sequences than T2-FLAIR. However, this study is limited to the in-plane motion artifact due to the fact T2-FLAIR images were acquired in 2D that is inherently contain geometry distortion along the slice directions.^36^

## 6 Conclusion

Our method, MAUDGAN, could disentangle motion artifacts from the 3D T1-Gd dataset under a multi-modal framework. The motion reduction will improve post-processing methods like manual and automatic brain tumors and organ at risk delineations and might increase the CT/MRI coregistration accuracy. Especially, the MAUDGAN would benefit elderly and infant patients with more involuntary motions during the 3D T1-Gd imaging with a long acquisition time. This retrospective motion correction is free from additional hardware or sequence modifications during the imaging, which makes it more practical.

## Data Availability

All data produced are available online at

https://wiki.cancerimagingarchive.net/pages/viewpage.action?pageId=95224486

## Acknowledgments

This work was supported by NSERC CREATE RHHDS program and NSERC discovery grant.

## Data Availability

The brain dataset was obtained from The Cancer Imaging Archieve (https://wiki.cancerimagingarchive.net/pages/viewpage.action?pageId=95224486).

## Footnotes

1 https://pytorch.org/

2 https://github.com/junyanz/pytorch-CycleGAN-and-pix2pix

